# Clinical Characteristics of 24 Asymptomatic Infections with COVID-19 Screened among Close Contacts in Nanjing, China

**DOI:** 10.1101/2020.02.20.20025619

**Authors:** Zhiliang Hu, Ci Song, Chuanjun Xu, Guangfu Jin, Yaling Chen, Xin Xu, Hongxia Ma, Wei Chen, Yuan Lin, Yishan Zheng, Jianming Wang, Zhibin Hu, Yongxiang Yi, Hongbing Shen

## Abstract

**Background:** Previous studies have showed clinical characteristics of patients with the 2019 novel coronavirus disease (COVID-19) and the evidence of person-to-person transmission. Limited data are available for asymptomatic infections. This study aims to present the clinical characteristics of 24 cases with asymptomatic infection screened from close contacts and to show the transmission potential of asymptomatic COVID-19 virus carriers.

**Methods:** Epidemiological investigations were conducted among all close contacts of COVID-19 patients (or suspected patients) in Nanjing, Jiangsu Province, China, from Jan 28 to Feb 9, 2020, both in clinic and in community. Asymptomatic carriers were laboratory-confirmed positive for the COVID-19 virus by testing the nucleic acid of the pharyngeal swab samples. Their clinical records, laboratory assessments, and chest CT scans were reviewed.

**Findings:** None of the 24 asymptomatic cases presented any obvious symptoms before nucleic acid screening. Five cases (20.8%) developed symptoms (fever, cough, fatigue, etc.) during hospitalization. Twelve (50.0%) cases showed typical CT images of ground-glass chest and 5 (20.8%) presented stripe shadowing in the lungs. The remaining 7 (29.2%) cases showed normal CT image and had no symptoms during hospitalization. These 7 cases were younger (median age: 14.0 years; *P* = 0.012) than the rest. None of the 24 cases developed severe COVID-19 pneumonia or died. The median communicable period, defined as the interval from the first day of positive nucleic acid tests to the first day of continuous negative tests, was 9.5 days (up to 21 days among the 24 asymptomatic cases). Through epidemiological investigation, we observed a typical asymptomatic transmission to the cohabiting family members, which even caused severe COVID-19 pneumonia.

**Interpretation:** The asymptomatic carriers identified from close contacts were prone to be mildly ill during hospitalization. However, the communicable period could be up to three weeks and the communicated patients could develop severe illness. These results highlighted the importance of close contact tracing and longitudinally surveillance via virus nucleic acid tests. Further isolation recommendation and continuous nucleic acid tests may also be recommended to the patients discharged.

## Introduction

The epidemic of the 2019 novel coronavirus disease (COVID-19) infections that developed in Wuhan, Hubei Province has spread to other provinces in China. As of 16 February 2020, 51, 857 patients with COVID-2019 have been reported and 1, 121 deaths were confirmed in China ^1^. To date, accumulated evidence has indicated person-to-person transmission both in hospital and family settings^2–6^. Since January 23, 2020, the source city was locked down to suppress the spread of the disease; however, the massive human movement during the traditional Chinese New Year holidays might have fueled the spread of the disease. At present, there were at least 10,000 cases in other provinces, mostly identified through symptoms plus recent travel history to Hubei province, China.

Genetic analysis of COVID-19 revealed that the virus was similar to severe acute respiratory syndrome coronavirus (SARS-CoV) ^7^. Unlike SARS-CoV, transmission of COVID-19 occurs during the prodromal period when those infected are mildly ill, and carry on usual activities, which contributes to the spread of infection ^8,9^. According to the report on “Diamond Princess”, among the 1,723 tested travelers, 189 asymptomatic individuals were positive for the COVID-19 virus as of 17 February 2020 ^10^, which indicated that a large number of asymptomatic carriers and mild patients remain undiscovered in the community. It is crucial to identify and isolate asymptomatic carriers and mild patients in order to contain the outbreaks in later stages.

Here, we conducted an epidemiological investigation among close contacts of COVID-19 patients in Nanjing, Jiangsu Province, China and identified 24 asymptomatic carriers. We sought to delineate the clinical characteristics and the transmission potential of asymptomatic infections.

## Methods

### Study design and patient

All screening-identified asymptomatic COVID-19 cases in Nanjing were admitted to the Second Hospital of Nanjing, the only hospital to admit for COVID-19 patients of the city, from Jan 28 to Feb 9, 2020. We retrospectively reviewed documents on epidemiological investigation and medical records. Asymptomatic carriers were screened from close contacts both in clinic and in community. Close contacts were defined as: 1) cohabiting family members of the COVID-19 patient or suspected patient; or 2) individuals who were exposed to the COVID-19 patient within 2 meters for more than 1 hour within 2 days before the symptom onset of the patient. Nucleic acid test was performed on the same day when the contacts were tracked. This study was reviewed and approved by the Medical Ethical Committee of Second Hospital of Nanjing (approval number 2020-LS-ky003). Written informed consent was obtained from each enrolled patient.

### Data source

Upon admission, detailed contact history of each case was collected, including their travel history in Hubei, the date/time of suspected contact, the symptom onset after contacting with the confirmed patient, and information of other family members. History of smoking and coexisting disorders was also collected.

For each case, computed tomography (CT) was performed upon admission. Laboratory assessments include whole blood count, blood chemistry, coagulation test, liver and renal function, electrolytes, C-reactive protein, procalcitonin, lactate dehydrogenase and creatine kinase. Pharyngeal swab specimens were collected on admission day and every other day thereafter for the COVID-19 virus test. All samples were processed simultaneously at the Department of Clinical Laboratory of Second Hospital of Nanjing. The symptoms presented on each case during their hospitalization were recorded in this study, including fever, cough, nasal congestion, dizziness, fatigue, arthralgia, etc. The virus clearance was defined as 2 continuous negatives of nucleic acid tests.

### Laboratory Nucleic Acid Test

Pharyngeal swab specimens, as described above, were tested for COVID-19 by using qRT-PCR kits (BGI Genomics, Beijing, China) recommended by the Chinese Center for Disease Control and Prevention (CDC) following WHO guidelines ^11^. The total nucleic acids were isolated from 200μl virus preservation solution containing throat swabs through automatic nucleic acid extraction system (BioPerfectus technologies company). The primers and probe set for open reading frame1ab (*ORF1 ab)* were as follows: forward primer 5’-AGAAGATTGGTTAGATGATGATAGT-3’; reverse primer 5’-TTCCATCTCTAATTGAGGTTGAACC-3’; and probe 5’-FAM-TCCTCACTGCCGTCTTGTTGACCA-BHQ1-3’. The human GAPDH gene was used as an internal control (forward primer 5’-TCAAGAAGGTGGTGAAGCAGG-3’; reverse primer 5’-CAGCGTCAAAGGTGGAGGAGT-3’; probe 5’-VIC-CCTCAAGGGCATCCTGGGCTACACTBHQ1-3’).

### Statistical analysis

Continuous variables were expressed as the medians and interquartile ranges (IQR). Categorical variables were summarized as the counts and percentages in each category. Wilcoxon rank-sum tests were applied to continuous variables, and Fisher’s exact tests were used for categorical variables. All analyses were conducted with STATA/SE 14.0.

## Result

### Demographic characteristics

The demographic characteristics of the 24 asymptomatic cases are shown in **Table S1**. None of the cases were healthcare workers and 8 (33.3%) had a history of recent travel to Hubei (Case 1 and 5 were residents of Hubei (marked with blue arrows), Case 3, 4, 6, 9, 13 and 17 have travelled to Hubei (marked with blue boxes), and the period in Hubei might be the suspected contact time. The suspected contact time of other cases who stay in Nanjing was marked with gray boxes according to the epidemiological investigation). The diagnosis date of each case showed that the number of cases who have been to Hubei decreased since Jan 28, 2020 (**Figure S1**). Individuals of all ages were involved in the COVID-19 asymptomatic infection with age ranging from 5 to 95 years old (median: 32.5 years) whereas 20.8% (5/24) of the cases were aged below 15 years. Eight cases (33.3%) were males. Two cases had a history of smoking (Case 1 and Case 13), and 2 were diagnosed with diabetes and hypertension (Case 8 and Case 13).

### Symptoms

Five cases (Case 2, 4, 5, 6, and 10, **Figure 1**) had symptoms during hospitalization. All the five cases developed fever without chills, with body temperatures fluctuating from 36.5°C to 38.0°C, but none presented high fever (body temperature >39°C). Cases 4, 6 and 10 were free of other symptoms. Case 2 also had cough, fatigue and nasal congestion, and Case 5 presented cough, fatigue, dizziness and arthralgia. Several cases also developed transient symptoms during hospitalization, including chills (Case 8), diarrhea (Case 21 and 22) and rashes (Case 16 and 18), that were discussed by clinical expert panel and considered as an infusion reaction of intravenous immunoglobulin, side effects of lopinavir/ritonavir and darunavir/cobicistat, respectively; and were therefore not grouped as cases with symptoms caused by COVID-19.

**Figure 1.**
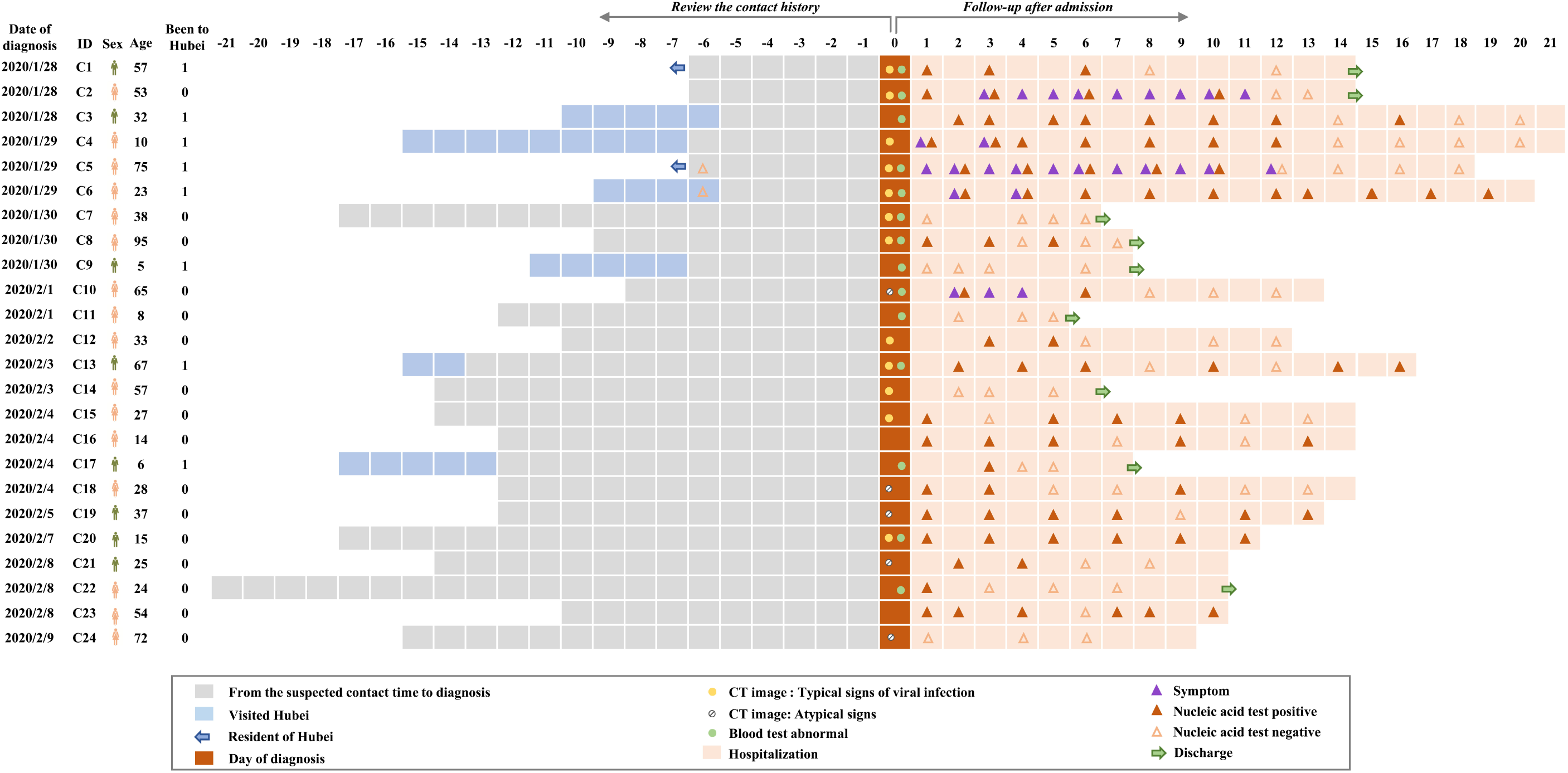
The epidemiological and clinical characteristic of the 24 asymptomatic infections with COVID-19 screened among close contacts. The cases were successively listed according to the date of diagnosis. Date of diagnosis was defined as origin point, the contact history was reviewed; abnormal blood test and CT image upon admission were listed; and symptom and nucleic acid test were recorded during hospitalization.

### Radiologic and laboratory findings

On admission, all 24 cases had a chest CT scan. Twelve (50.0%) cases showed typical findings of chest CT images of ground-glass or patchy shadows in the lungs (**Figure S2**). Five (20.8%) cases showed stripe shadows in lungs, an atypical image finding. CT scans of the rest 7 (29.2%) cases showed normal images. Four (16.7%) of the 24 cases had lymphopenia (<0.8 × 10^9^ cells/L) on admission. Leukopenia was also observed in 4 cases. Elevated levels of alanine aminotransferase, aspartate aminotransferase, creatine kinase, C-reactive protein and D-dimer were uncommon (**Table S1**). During hospitalization, Case 2 and Case 5 developed leukopenia. Seven cases presented an elevated level of serum lactose dehydrogenase, among whom 3 were accompanied with an increased level of C-reactive protein (**Figure S3**).

### Treatment and clinical outcome

Antiviral therapy was given to 21 cases (87.5%) as initiated therapy, in which, one case also received antibiotics therapy, antifungal therapy plus immunoglobin therapy; and immunoglobin therapy was also given to 2 cases. All these cases were treated with interferon atomization. None of the cases developed severe pneumonia, requiring systemic corticosteroids treatment, mechanical ventilation, or admission to ICU, and none died (**Table S1**). As of Feb 18, 2020, a total of 18 cases (75.0%) had the virus cleared (2 continuous negatives of nucleic acid tests), among whom 9 cases were discharged from the hospital while the rest 9 were kept in hospital for further observation. Six cases (Case 3, 8, 13, 16, 19 and 23) had nucleic acid tests reversed to positive after one negative result. Of particular concern, Case18 showed positive again even after the continuous negative of nucleic acid tests. Five cases (Case 7, 9, 11, 14 and 24) had the virus cleared within a short time. The communicable period, defined as the interval from the first day of positive nucleic acid tests to the first day of continuous negative tests, ranged from 1 to 21 days (median: 9.5 days, IQR, 3.5-13.0 days, **Figure S4, Table S1**). Due to the censored data of 6 cases and the indeterminate date of the exact first infection, the communicable period was underestimated.

### The characteristics of asymptomatic plus CT normal patients

We found that 7 (29.2%) cases had normal CT images and no symptoms during hospitalization. As compared with other cases, these 7 cases were younger (median, 14.0; *P* = 0.012) (**Table S2)**. There were no differences in other characteristics between these 2 groups, possibly due to the limited sample size. The blood test of these 7 cases showed no obvious abnormity. The median communicable period of the 7 patients was 4.0 days (IQR, 2.0-15.0).

### The transmission evidence of the asymptomatic COVID-19 carrier - the family-based transmission of Case 13

We reviewed the medical records and epidemiological history of each case and of their family members if they were admitted in hospital for confirmed or suspected COVID-19 infection. In the family of Case 13, Relative 1 (wife of Case 13) first visited the hospital (**Figure 2**). Relative 1, a 64-year-old otherwise healthy woman, developed fever (the highest temperature was 38.7°C), cough, fatigue and vomiting on Jan 30, 2020. Three days later (Feb 2, 2020), she visited the hospital and was tested positive for the COVID-19 virus. Then, epidemiological investigations and nucleic acid tests were conducted on her son (Relative 2), daughter-in-law (Relative 3) and Case 13. Relative 2 and 3 had developed respiratory symptoms before investigation and were demonstrated positive for the COVID-19 virus. Nucleic acid test was also positive for Case 13, but he had no symptoms during admission. Through epidemiological investigation, Case 13 admitted his travel history to the city of Huanggang, Hubei province on Jan 19-20, 2020. Relative 1, 2 and 3 are all local Nanjing residents and denied the history of contact with any confirmed or suspected COVID-19 patient, except for Case 13, in the recent 14 days. During hospitalization, Case 13 had no symptoms but remained positive for the COVID-19 virus till Feb 18, 2020, which indicated that the communicable period could be as long as 29 days (from 21 Jan to 18 Feb, 2020). Among the 3 infected family members, Relative 1 developed severe COVID-19 pneumonia and was admitted to the ICU on Feb 6, 2020. Relative 2 and 3 demonstrated clearance of the virus on Feb 18, 2020. In brief, Case 13, an asymptomatic COVID-19 carriers transmitted the virus to his cohabiting family members, and 1 of the infected individuals developed severe COVID-19 pneumonia and was admitted to ICU.

**Figure 2.**
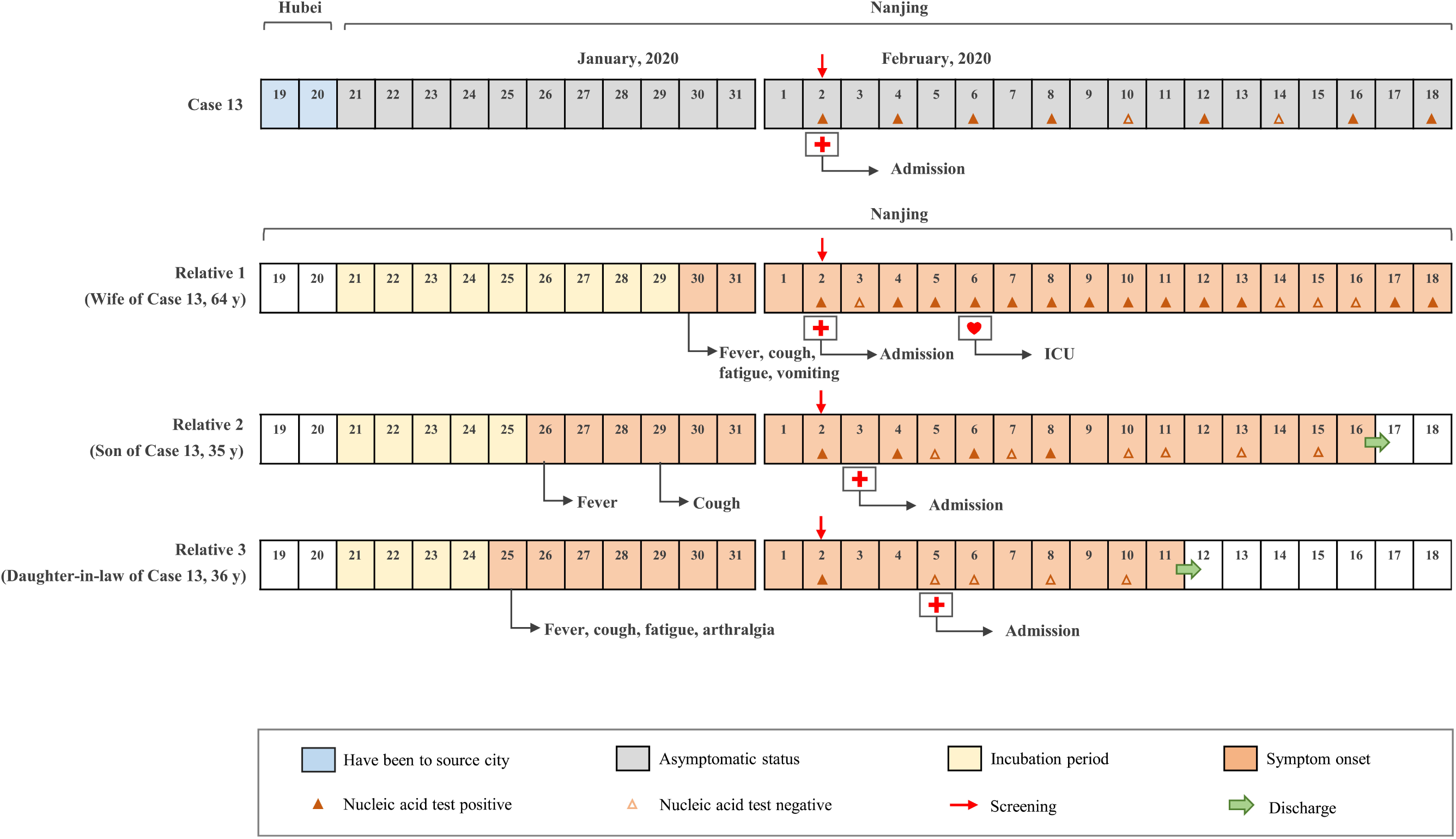
The transmission evidence from an asymptomatic COVID-19 carrier to his cohabited family members, and his wife admitted to the ICU after being infected.

## Discussion

In the present study, we provided epidemiological and clinical data of 24 asymptomatic COVID-19 infections identified from the screening of close contacts in Nanjing, Jiangsu Province. These asymptomatic cases were mildly ill as compared to those previously reported in Wuhan, Hubei^5,12,13^. None of the 24 cases developed severe pneumonia as of Feb18, 2020, and only 5 cases showed typical symptoms during hospitalization. Similar to previous studies^13,14^, fever, cough and fatigue were the main symptoms. Lymphopenia and leukopenia, previously associated with disease severity ^5,13,15^, were uncommon in the asymptomatic cases of the present study. Interestingly, we found that young cases (< 15 years old) were prone to be asymptomatic even during hospitalization and to have a normal CT image, which partially suggested that nucleic acid testing would be crucial to identify asymptomatic infections in young close contacts.

Notably, we provided evidence for transmission from an asymptomatic infector to close contacts that led to severe COVID-19 pneumonia. These findings indicate that asymptomatic carriers can result in person-to-person transmission and should be considered a source of COVID-19 infection. It is also implicated by the report of the “Diamond Princess” that large numbers of asymptomatic or mild patients might have been hiding in the community. Therefore, it is of great public health significance to strictly monitor close contacts via multiple nucleic acid screenings to contain potential outbreaks. Of particular, since massive movement returning to the work place and the school is in progress, active contact tracing and strict health monitoring should remain an important strategy in China, or worldwide. Guidance of self-protection, active isolation of close contacts (either at home or centralized) should be continuously highlighted. Isolation and multiple virus nucleic acid detections are also recommended for discharged COVID-19 patients.

Our study was conducted in Nanjing, a city with adequate medications and public health interventions. The included cases were identified from systematic screening of the close contacts of COVID-19 patients (or suspected patients) both in clinic and in community. All asymptomatic carriers were admitted to and treated in the Second Hospital of Nanjing, and therefore the study population has strong representativeness of the whole city. We collected the contact history and followed longitudinal nucleic acid tests of the close contacts during their hospitalization, which provided a relatively reliable communicable period of asymptomatic cases. However, it is notable that the real communicable period was longer than was calculated, due to the censored data of 6 patients and the indeterminate date of the exact first infection. This study is limited by the small sample size and lack of data on nucleic acid tests before the diagnosis date. Large-scale multicenter studies are needed to verify our findings.

## Data Availability

We obtained the data from the Second Hospital of Nanjing. The ownership of the data belongs to the Second Hospital of Nanjing. Researchers who meet the criteria for access to confidential data can contact the at the the Second Hospital of Nanjing to request the data.

## Figure Legend

**Figure S1. Date of asymptomatic COVID-19 infection onset among close contact in Nanjing and the distribution of travel history to Hubei**.

**Figure S2. Representative chest CT abnormalities of asymptomatic COVID-19 infections. Case 4: Chest CT upon admission showed small local ground glass opacity (A, arrow), which completely resolved after 13 days (B). Case 13: Chest CT upon admission showed multiple ground glass opacities (C, arrow). A Chest CT 8 days later showed substantial resolution of those lesions (D). Case 18: Chest CT upon admission showed a small fibrous strip (E, arrow), which was stable on a follow-up chest CT after 4 days**.

**Figure S3. The abnormal blood test during hospitalization**.

**Figure S4. The calculation process of the communicable period**.

## Acknowledgement

This study was funded in part by the project of Jiangsu province medical youth talent (QNRC2016059), Nanjing medical science and technique development foundation (ZKX17040 and YKK18153), the National Natural Science Foundation of China (81903382), Cheung Kong Scholars Program of China. Written informed consent was obtained from each enrolled patient.

## Author Approval

All authors have seen and approved the manuscript.

## Data sharing

With the permission of the corresponding authors, we can provide participant data without names and identifiers, but not the study protocol, statistical analysis plan, or informed consent form. Data can be provided after the Article is published. Once the data can be made public, the research team will provide an email address for communication. The corresponding authors have the right to decide whether to share the data or not based on the research objectives and plan provided.

## Ethical approval

This study was reviewed and approved by the Medical Ethical Committee of Second Hospital of Nanjing (approval number 2020-LS-ky003).

## Author’s contribution

Yongxiang Yi, Zhibin Hu, Zhiliang Hu and Guangfu Jin made substantial contributions to the study concept and design. Yongxiang Yi and Zhiliang Hu took responsibility for the acquisition and check of data. Zhibin Hu, Zhiliang Hu, Ci Song took responsibility for analysis and interpretation of the data; Wei Chen was responsible for laboratory detection; Ci Song and Zhibin Hu were in charge of the manuscript draft. Ci Song and Xin Xu took responsibility for the statistical analysis; Chuanjun Xu, Yaling Chen, Yishan Zheng and Jianming Wang provided administrative, technical, and material support; Yongxiang Yi, Zhibin Hu and Hongbing Shen made substantial revisions to the manuscript.

